# Artificial Intelligence for context-aware surgical guidance in complex robot-assisted oncological procedures: An exploratory feasibility study

**DOI:** 10.1101/2022.05.02.22274561

**Authors:** Fiona R. Kolbinger, Sebastian Bodenstedt, Matthias Carstens, Stefan Leger, Stefanie Krell, Franziska M. Rinner, Thomas P. Nielen, Johanna Kirchberg, Johannes Fritzmann, Jürgen Weitz, Marius Distler, Stefanie Speidel

## Abstract

**Introduction:** Complex oncological procedures pose various surgical challenges including dissection in distinct tissue planes and preservation of vulnerable anatomical structures throughout different surgical phases. In rectal surgery, violation of dissection planes increases the risk of local recurrence and autonomous nerve damage resulting in incontinence and sexual dysfunction. This work explores the feasibility of phase recognition and target structure segmentation in robot-assisted rectal resection (RARR) using machine learning.

**Materials and Methods:** A total of 57 RARR were recorded and annotated with respect to surgical phases and exact locations of target structures (anatomical structures, tissue types, static structures, and dissection areas). For surgical phase recognition, three machine learning models were trained: LSTM, MSTCN, and TransSVNet. Based on pixel-wise annotations of target structures in 9037 images, individual segmentation models based on DeepLabV3 were trained. Model performance was evaluated using F1 score, Intersection-over-Union (IoU), accuracy, precision, recall, and specificity.

**Results:** The best results for phase recognition were achieved with the MSTCN model (F1 score: 0.82 ± 0.01, accuracy: 0.84 ± 0.03). Mean IoUs for target structure segmentation ranged from 0.14 ± 0.22 to 0.80 ± 0.14 for organs and tissue types and from 0.11 ± 0.11 to 0.44 ± 0.30 for dissection areas. Image quality, distorting factors (i.e. blood, smoke), and technical challenges (i.e. lack of depth perception) considerably impacted segmentation performance.

**Conclusion:** Machine learning-based phase recognition and segmentation of selected target structures are feasible in RARR. In the future, such functionalities could be integrated into a context-aware surgical guidance system for rectal surgery.

## 1 Introduction

Despite advances in nonsurgical management of rectal cancer, the majority of curative treatment approaches require oncologically radical surgical removal of the primary tumor and locoregional lymph nodes within the mesorectal envelope (total mesorectal excision, TME). This surgical step requires delicate preparation along the mesorectal fascia to ensure complete resection and minimize the risk of local tumor recurrence [1,2]. In parallel, autonomous nerves running in close proximity to the resection area must be protected to avoid postoperative incontinence and sexual dysfunction. With highly varying numbers depending on surgeon expertise and center volume, these functional sequelae affect up to 85% of patients after oncological rectal resection and can be a direct result of suboptimal surgical preparation outside of anatomical planes, leading to nerve injury [3,4].

Surgical robots offer mechanical enhancements such as improved instrument dexterity, hand-eye coordination, and a three-dimensional view, however in many types of surgery including rectal resection, no robust clinical benefit over conventional laparoscopic surgery could be demonstrated to date [5]. Based on the hypothesis that an integration of surgical guidance (i.e. segmentation overlays of specific structures in specific phases of a procedure) into minimally-invasive surgical systems could markedly improve surgical quality and outcome and diminish quality variation between surgeons, extensive research on the development of image recognition algorithms has been conducted in recent years using laparoscopic imaging data. Most of these endeavors, however, have focused on tasks with indirect surgical benefit, such as automated instrument detection [6,7]. Thus far, translational Artificial Intelligence (AI)-based success stories in the field of surgery are lacking and clinical applications are mostly limited to orthopedic, neurosurgical, and hepatic surgical procedures [8,9]. With regard to approaches with high translational potential in laparoscopic surgery, deep learning-based algorithms have recently been shown to identify relevant anatomical areas during cholecystectomy [10,11] and organs during laparoscopy [12,13]. Of note, such methods have primarily been established for less complex surgical procedures, for which – in part – open-access datasets exist for the purpose of algorithm development and validation [14,15]. In these operations, however, the benefit of AI-based assistance is marginal. In this project, we explore the potential of deep learning-based phase recognition – a prerequisite for context awareness – and target structure segmentation. To this end, we applied algorithms based on state-of-the-art convolutional neural networks (CNNs) to a dataset of spatio-temporal data in the form of temporally annotated robot-assisted rectal resection (RARR) videos and semantically segmented video frames. In the future, automatic phase recognition and segmentation of relevant anatomical structures could serve as a basis for a context-aware surgical guidance system for rectal surgery.

## 2 Material and Methods

### 2.1 Patient population and dataset

Between February 2019 and January 2021, video data from 57 robot-assisted anterior rectal resections, intersphincteric resections, or abdominoperineal resections were prospectively collected at the University Hospital Carl Gustav Carus Dresden, Germany (Appendix A). All patients had a clinical indication for the surgical procedure, recommended by an interdisciplinary tumor board. Procedures were performed using the da Vinci® Xi system (Intuitive Surgical, Sunnyvale, CA, USA). Surgeries were recorded using the CAST system (Orpheus Medical GmBH, Frankfurt a.M., Germany) and saved at a resolution of 1920 x 1080 pixels in MPEG-4 format.

This study was performed in accord with the ethical standards of the Helsinki declaration and its later amendments. The local Institutional Review Board (ethics committee at the Technical University Dresden) reviewed and approved this study (approval number: BO-EK-140032021). Written informed consent was waived. The trial was registered on clinicaltrials.gov (trial registration ID: NCT05268432).

### 2.2 Temporal annotation

For establishment of the phase recognition models, the dataset of 57 consecutive RARR recordings was randomly split into a dataset for model pre-training (24 temporally non-annotated videos) and a dataset for model training and validation (33 temporally annotated videos, Figure 1). According to a previously created protocol (Appendix B), the surgical process was temporally annotated using b<>com *Surgery Workflow Toolbox* [Annotate] version 2.2.0 (b<>com, Cesson-Sévigné, France) (Figure 1). The definition and order of the five phases – preparation and intraabdominal orientation, medial mobilization of descending colon, lateral mobilization of descending colon, mesorectal excision, and extraabdominal preparation – was based on institutional standards and international recommendations for RARR [16,17]. Splenic flexure and descending colon mobilization was carried out from medial to lateral as described previously by Ahmed *et al.* [16].

**Figure 1:**
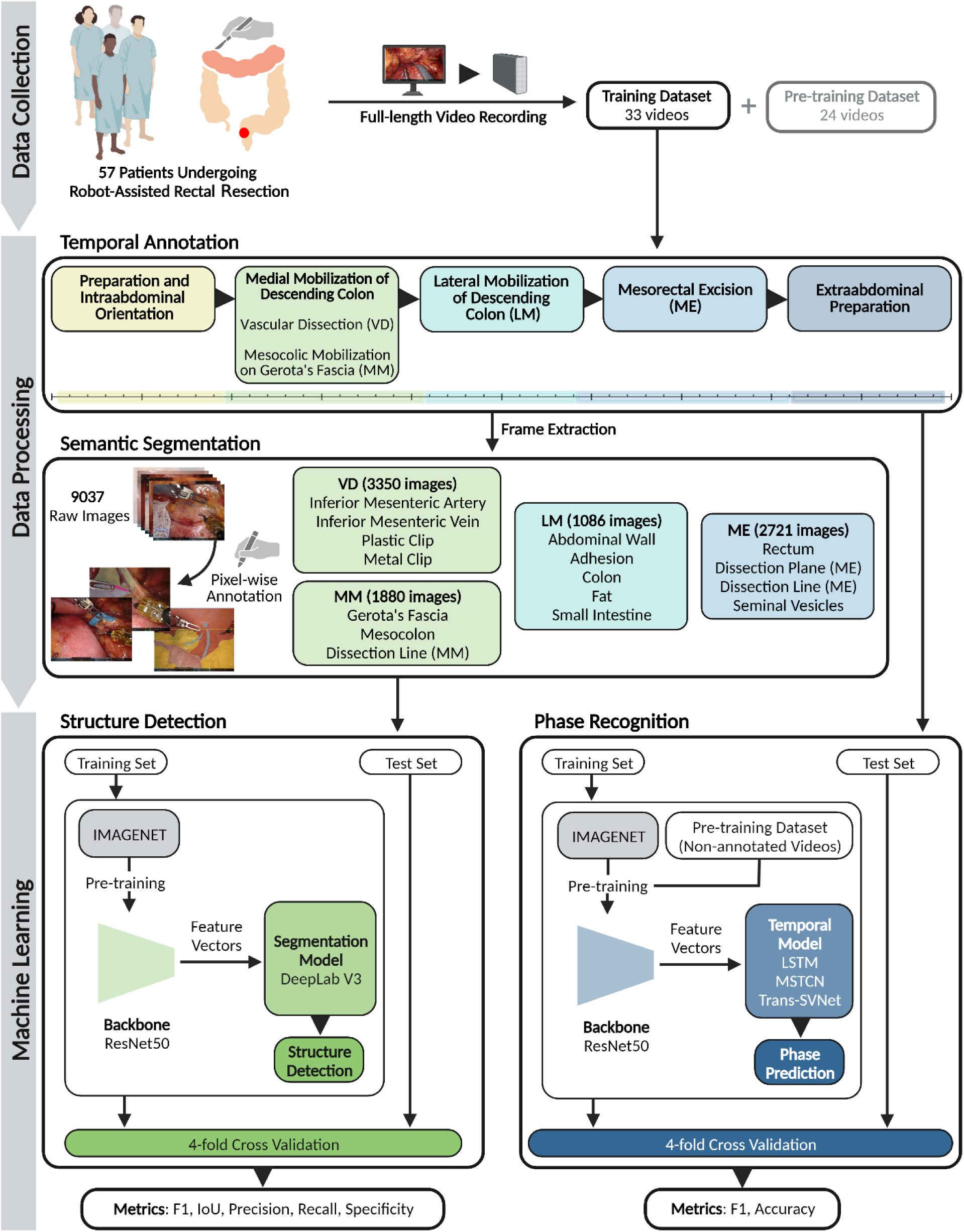
Overview of the data collection, data processing, and machine learning process in this study. Robot-assisted rectal resections were recorded and video data was annotated with respect to surgical phases and exact location of target structures. Phase recognition and structure segmentation models were based on a ResNet50 as backbone and combined with temporal models or segmentation models, respectively. Model performance was assessed using 4-fold cross validation and classical computer vision metrics. Abbreviations: Intersection over union (IoU), Lateral mobilization (LM), Medial mobilization (MM), Mesorectal excision (ME), Vascular dissection (VD).

### 2.3 Semantic segmentation

Phase-specific semantic segmentation algorithms were trained and validated on image data from the 33 temporally annotated surgery videos (Figure 1). For segmentation model training and validation, equidistant frames from at least ten selected video recordings for each phase were selected based on criteria of target structure visibility and exposition (Appendix C) and annotated with regard to the exact localization of anatomical structures and dissection planes (Figure 1, Appendix D, Appendix E) using the open-source software 3D Slicer with the SlicerRT program extension [18]. A detailed annotation protocol is provided in Appendix D. Annotations were performed by two fifth-year medical students (MC, FMR) after thorough theoretical and practical education in macroscopic and surgical anatomy and several rounds of training annotations with two experts in robot-assisted rectal surgery (JW, MD), and reviewed by a surgeon in training with four years of experience in robot-assisted rectal surgery (FRK).

### 2.3 Model development and evaluation: Phase recognition

For identification of the five surgical phases, we applied a typical two-stage pipeline. In the first stage, a residual neural network (ResNet50) [19] was used to learn image features, having demonstrated a high performance on the Cholec80 dataset [20]. In the second stage, the features of the ResNet50 were used to learn the temporal context with the following models: standard long short-term memory network (LSTM) [21], MSTCN [20], and Trans-SVNet [22] (Figure 1). Trans-SVNet has a third step, in which the features of ResNet50 and the features of MSTCN are used to train a Transformer model.

Pre-training of the feature extractor was performed in two different configurations: (a) the ResNet50 was pre-trained on the ImageNet database [23] only and (b) the pre-trained feature extractor was fine-tuned on 24 randomly selected temporally non-annotated RARR videos using self-supervised training. The self-supervised learning approach described by Funke *et al*. [24] was adapted, so that a ResNet50 was trained based on temporally coherent video frame embedding using 1st and 2nd order contrastive loss [25,26].

Afterwards, the two networks were trained frame-wise with the label information of the current phase. Data augmentation consisted of image rotation, flipping and noise. The network was trained for 30 epochs, each consisting of 3000 random chosen batches with a size of 64. As optimizer, AdamW [27] with an initial learning rate of 0.0001 was used and the weighted cross entropy loss was applied to handle the class-imbalance problem.

As self-supervised pre-training did not result in any performance benefits (Appendix F), the ResNet50 pre-trained on ImageNet was selected as feature extractor and combined with the temporal models. The 33 temporally annotated RARR videos were used for training and evaluated within a 4-fold cross validation scheme (Figure 1). We strictly avoided data leakage by splitting the folds for cross validation along patient lines, i.e. if videos from patient A were included in the training data, no videos from patient A were used for testing. Performance was evaluated on the test folds using F1 score and accuracy. These metrics were first computed per surgery video and then averaged over all patients.

### 2.4 Model development and evaluation: Semantic segmentation

For training of the segmentation algorithms for each individual phase, at least ten video recordings were selected per phase with regard to visibility of anatomical structures and dissection planes (Appendix D) and annotated as described above. To account for the differential visibility and relevance of individual segments, a separate convolutional neural network was trained for each phase for the phase-specific segmentation of target structures and tissue planes. For this purpose, the DeepLabv3 [28] framework was used with a ResNet50 backbone with default PyTorch pretraining on the COCO dataset [29]. The model was trained for 100 epochs with a batch size of 16 and evaluated within a 4-fold cross validation scheme (Figure 1). Data augmentation consisting of image resizing, image shifts and rotations was used to enhance the training process. For optimization of the network, the AdamW algorithm [27] with a learning rate of 0.0001 was used. The final model for each training fold was selected after 100 training iterations. Training parameters were selected for convergence of the loss on the training dataset. We strictly avoided data leakage by splitting the folds for cross validation along patient lines, i.e. if frames from patient A were included in the training data, no frames from patient A were used for testing. Segmentation performance was assessed on the test folds using F1 score, IoU, precision, recall, and specificity [30], commonly used technical measures of prediction exactness, ranging from 0 (least exact prediction) to 1 (entirely correct prediction). Segmentation metrics were summarized using mean and standard deviation. We provide means and standard deviations averaged both over all frames of a segment (irrespective of the surgery the frames originated from) and over average metrics of a segment from one surgery (i.e. averaging over frames from one surgery first, then averaging the surgery averages). In addition, the results of the semantic segmentation model were assessed qualitatively, comparing results for different structures and for individual images, to identify common challenges of segmentation in laparoscopy.

## 3 RESULTS

### 3.1 Dataset

In total, 57 RARR with an average duration of 414 ± 112 minutes performed by nine experts in robot-assisted oncological surgery were recorded and analyzed. The majority of patients (n = 43, 75.4%) were male, the predominant indication for the surgical procedure was colorectal cancer (n = 54, 94.7%). Twenty-five patients (43.9%) had undergone neoadjuvant irradiation to the surgical field. Overall, the sample represents a wide variety of rectal cancer manifestations, including a considerable number of patients (n = 27, 47.4%) with locally advanced tumors (Figure 1, Appendix A).

For training of surgical phase recognition models, 33 videos were temporally annotated, while 24 non-annotated videos were used for self-supervised pre-training (Figure 1). A distribution of phase durations is provided in Appendix G.

Individual segmentation models were trained for four relevant phases of RARR: Vascular Dissection (VD), Medial Mobilization of Descending Colon (MM), Lateral Mobilization of Descending Colon (LM), and Mesorectal Excision (ME). Distinct target structures were segmented in images from at least 10 patients per phase-specific segmentation model (Figure 1). Appendix E displays the number of annotated images per segment, while a distribution of respective segment sizes is provided in Appendix H.

### 3.2 Surgical phase recognition

Thirty-three videos were temporally annotated with regard to the sequence of five standardized surgical phases (Figure 1). The most accurate phase recognition results were achieved with MSTCN [20] (F1 score: 0.82 ± 0.01), outperforming the standard LSTM [21] (F1 score: 0.79 ± 0.02) and Trans-SVNet [22] (F1 score: 0.79 ± 0.01) (Table 1).

**Table 1:** Summary of performance metrics for surgical phase recognition using a standard LSTM, MSTCN, and Trans-SVNet. For each metric, mean and standard deviation are displayed.

| Temporal Model | F1 score | Accuracy |
| --- | --- | --- |
| LSTM | $0.79 \pm 0.02$ | $0.82 \pm 0.01$ |
| MSTCN | **$0.82 \pm 0.01$** | **$0.84 \pm 0.03$** |
| Trans-SVNet | $0.79 \pm 0.01$ | $0.83 \pm 0.02$ |

### 3.3 Segmentation of anatomical structures

For semantic segmentation, individual models based on DeepLabv3 [28] were trained to identify 16 distinct target structures, encompassing nine anatomical structures (Inferior Mesenteric Artery, Inferior Mesenteric Vein, Gerota’s fascia, the mesocolon, the abdominal wall, the colon, the small intestine, the rectum, and seminal vesicles), two additional tissue types (adhesion and fat), two static structures (plastic clip and metal clip), and three dissection areas (the dissection line during medial mobilization of the colon, the avascular dissection plane as well as the exact dissection line during mesorectal excision). In total, 9037 annotated frames were used for training of the semantic segmentation algorithms (Figure 1, Appendix D, Appendix E). Table 2 displays mean segmentation metrics for individual segments predicted by the trained phase-specific algorithms and averaged over all frames irrespective of the surgery the frames originated from. The performance metrics were overall very similar when computed over segment averages of each surgery (Appendix I).

**Table 2:** Summary of performance metrics for phase-specific semantic segmentation of target structures and dissection areas. For each metric, mean and standard deviation are displayed. Means and standard deviations were calculated over metrics of each individual image. Abbreviations: Intersection over union (IoU), Lateral mobilization (LM), Medial mobilization (MM), Mesorectal excision (ME), Vascular dissection (VD).

| Phase | Target Structure | F1 score | IoU | Precision | Recall | Specificity |
| --- | --- | --- | --- | --- | --- | --- |
| VD | Inferior Mesenteric Artery | $0.29 \pm 0.30$ | $0.21 \pm 0.24$ | $0.48 \pm 0.41$ | $0.36 \pm 0.30$ | $1.00 \pm 0.01$ |
| | Inferior Mesenteric Vein | $0.33 \pm 0.34$ | $0.26 \pm 0.28$ | $0.47 \pm 0.39$ | $0.48 \pm 0.37$ | $1.00 \pm 0.00$ |
| | Plastic Clip | $0.35 \pm 0.38$ | $0.28 \pm 0.32$ | $0.54 \pm 0.33$ | $0.47 \pm 0.45$ | $1.00 \pm 0.00$ |
| | Metal Clip | $0.11 \pm 0.21$ | $0.07 \pm 0.15$ | $0.12 \pm 0.21$ | $0.47 \pm 0.42$ | $1.00 \pm 0.01$ |
| MM | Gerota's Fascia | $0.84 \pm 0.18$ | $0.75 \pm 0.21$ | $0.82 \pm 0.21$ | $0.90 \pm 0.12$ | $0.94 \pm 0.06$ |
| | Mesocolon | $0.79 \pm 0.20$ | $0.69 \pm 0.23$ | $0.79 \pm 0.24$ | $0.84 \pm 0.16$ | $0.94 \pm 0.07$ |
| | Dissection Line (MM) | $0.21 \pm 0.21$ | $0.13 \pm 0.14$ | $0.21 \pm 0.22$ | $0.39 \pm 0.29$ | $1.00 \pm 0.00$ |
| LM | Abdominal Wall | $0.88 \pm 0.10$ | $0.80 \pm 0.14$ | $0.88 \pm 0.13$ | $0.90 \pm 0.10$ | $0.96 \pm 0.04$ |
| | Adhesion | $0.33 \pm 0.27$ | $0.23 \pm 0.20$ | $0.37 \pm 0.31$ | $0.47 \pm 0.27$ | $0.99 \pm 0.01$ |
| | Colon | $0.62 \pm 0.35$ | $0.53 \pm 0.33$ | $0.67 \pm 0.36$ | $0.74 \pm 0.27$ | $0.98 \pm 0.02$ |
| | Fat | $0.79 \pm 0.22$ | $0.69 \pm 0.23$ | $0.78 \pm 0.24$ | $0.87 \pm 0.14$ | $0.95 \pm 0.05$ |
| | Small Intestine | $0.26 \pm 0.38$ | $0.22 \pm 0.33$ | $0.33 \pm 0.43$ | $0.57 \pm 0.38$ | $1.00 \pm 0.01$ |
| ME | Rectum | $0.51 \pm 0.27$ | $0.38 \pm 0.23$ | $0.54 \pm 0.29$ | $0.64 \pm 0.26$ | $0.99 \pm 0.03$ |
| | Dissection Plane (ME) | $0.54 \pm 0.33$ | $0.44 \pm 0.30$ | $0.59 \pm 0.34$ | $0.63 \pm 0.32$ | $0.94 \pm 0.06$ |
| | Dissection Line (ME) | $0.17 \pm 0.17$ | $0.11 \pm 0.11$ | $0.17 \pm 0.17$ | $0.25 \pm 0.23$ | $1.00 \pm 0.00$ |
| | Seminal Vesicles | $0.18 \pm 0.29$ | $0.14 \pm 0.22$ | $0.30 \pm 0.38$ | $0.33 \pm 0.35$ | $1.00 \pm 0.00$ |

Overall, segmentation model performance varied strongly both between structures (mirrored by variations in mean performance metrics) and between individual images (mirrored by large standard deviations and uneven metric distributions within one structure) (Table 2, Figure 2, Figure 3). Among anatomical structures, segmentation performance was highest for the abdominal wall (F1 score: 0.88 ± 0.10), Gerota’s fascia (F1 score: 0.84 ± 0.18), and the mesocolon (F1 score: 0.79 ± 0.20), whereas segmentation performance was weaker for seminal vesicles (F1 score: 0.18 ± 0.29), the small intestine (F1 score: 0.26 ± 0.38), and intestinal vessels (F1 score: 0.29 ± 0.30 for the Inferior Mesenteric Artery and 0.33 ± 0.34 for the Inferior Mesenteric Vein) (Table 2, Figure 2).

**Figure 2:**
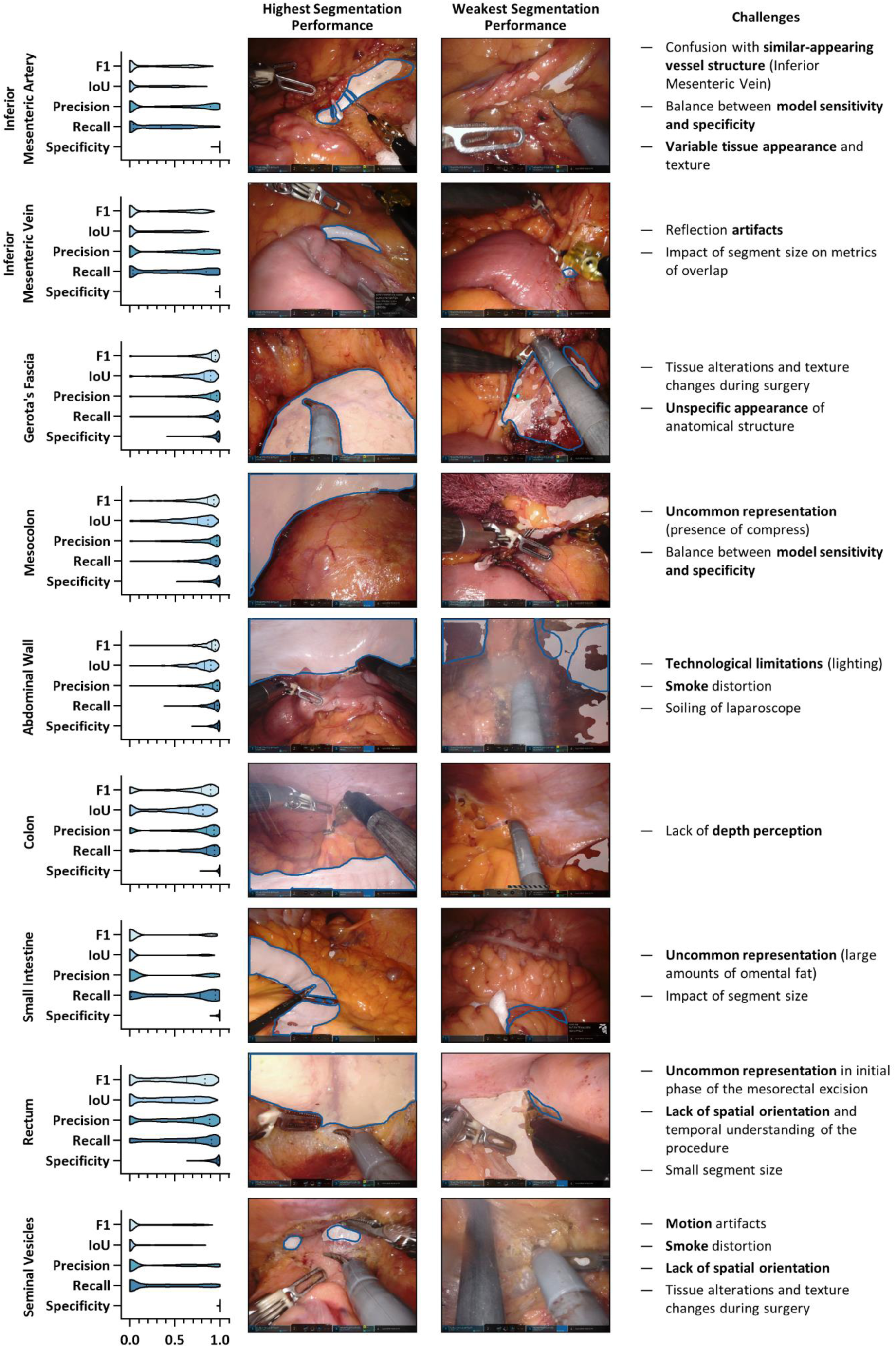
Performance of machine learning models based on DeepLabv3 for segmentation of anatomical structures. Violin plot illustrations display the distributions of performance metrics, with the median and quartiles illustrated as solid and dashed lines, respectively. Example images with the highest and weakest segmentation performance (by IoU) are displayed for each target structure. Ground truth is displayed as blue line, model segmentations are displayed as white overlay. Abbreviations: F1 score (F1), Intersection over union (IoU).

**Figure 3:**
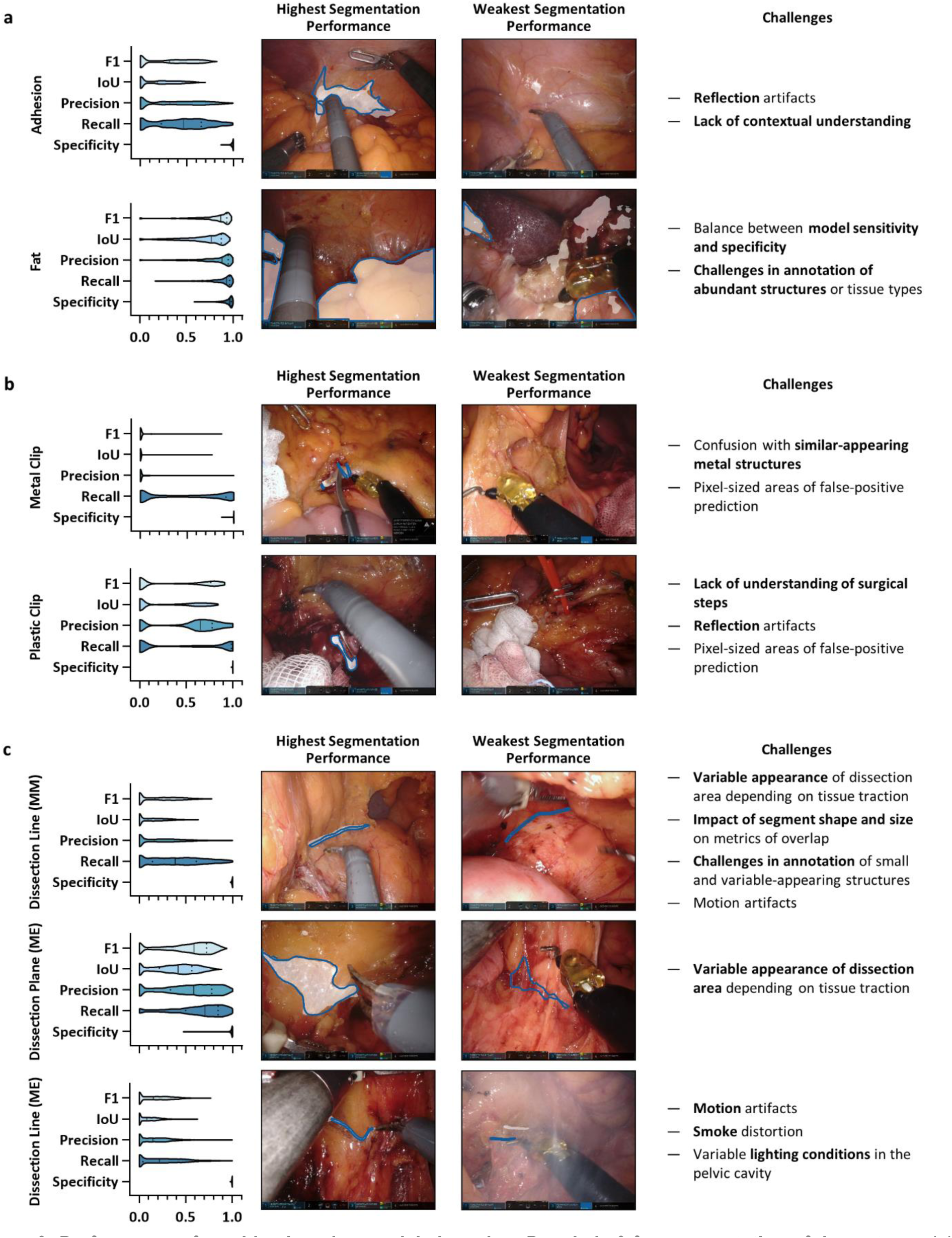
Performance of machine learning models based on DeepLabv3 for segmentation of tissue types (a), static structures (b), and dissection areas (c). Violin plot illustrations display the distributions of performance metrics, with the median and quartiles illustrated as solid and dashed lines, respectively. Example images with the highest and weakest segmentation performance (by IoU) are displayed for each target structure. Ground truth is displayed as blue line, model segmentations are displayed as white overlay. Abbreviations: F1 score (F1), Intersection over union (IoU), Medial Mobilization phase (MM), Mesorectal Excision phase (ME).

### 3.4 Segmentation of tissue types, static structures and dissection planes

To explore the capability to provide spatial information exceeding anatomical structures, segmentation models based on DeepLabv3 were trained to delineate tissue types with relevance in the context of surgical challenges (fat and adhesion), surgical clips that often require accurate positioning, and dissection areas that are of prognostic relevance in rectal surgery.

Fat was among the best-predicted structures overall (F1 score: 0.79 ± 0.22), while prediction quality of tissue adhesion was variable and overall weaker than several organs (F1 score: 0.33 ± 0.27) (Table 2, Figure 2, Figure 3 a). The plastic and the metal clip, both small but distinct-appearing structures, were falsely not predicted in several images displaying them, resulting in a large proportion of unpredicted images (Figure 3 b). During mesorectal excision, the avascular dissection plane could be predicted with an F1 score of 0.54 ± 0.33, whereas prediction of the exact dissection lines both during medial mobilization of the mesocolon and mesorectal dissection was largely unsuccessful and, when predicted, highly variable at F1 scores of 0.21 ± 0.21 and 0.17 ± 0.17, respectively (Table 2, Figure 3 c).

### 3.5 Challenges of segmentation in laparoscopy

Review of all segmentation results including images with high and weak segmentation performance revealed several common technological and methodological challenges related to segmentation algorithms that limit model performance and clinical impact (Figure 2, Figure 3, Figure 4). First, segment size substantially impacted segmentation performance, with a trend towards better prediction of large, uniform-appearing organs such as the abdominal wall or the mesocolon as compared to small or tubular and variable-appearing structures such as seminal vesicles and dissection planes. In turn, some larger structures such as Gerota’s fascia, the mesocolon, or fat were prone to unspecific false positive prediction (Figure 2, Figure 3). Second, images distorted by excessive amounts of blood, smoke, soiling of the endoscope lens, or blurring caused by camera shake tended to result in false-negative predictions and overall inferior segmentation performance (Figure 2, Figure 3). At the same time, surgery-induced changes to the surgical field that alter the appearance of anatomical structures have the potential to create unspecific visual appearances of organ surfaces, leading to both false-positive and false-negative outputs. Third, there were specific prediction errors for selected target structures; for example, the metal clip was occasionally predicted as part of surgical instruments and the mesenteric vessels (inferior mesenteric artery and vein) were frequently confused (Figure 2, Figure 3).

**Figure 4:**
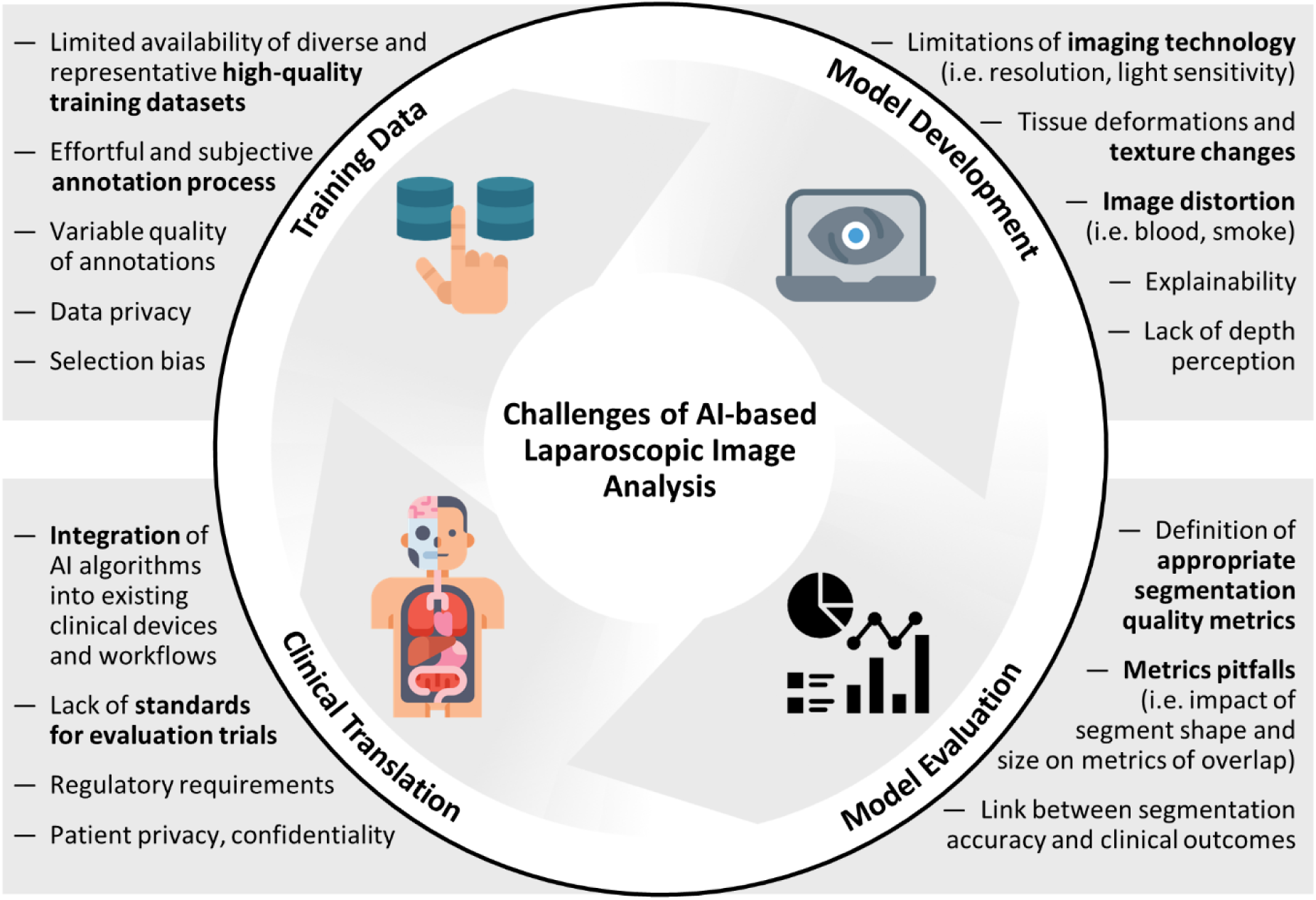
Challenges of AI-based laparoscopic image analysis. This figure highlights existing challenges of target structure segmentation in laparoscopic imaging related to training data, model development, model evaluation, and clinical translation.

## 4 Discussion

Correct recognition of visual cues is a key challenge in minimally-invasive surgery, as the majority of currently available laparoscopic and robotic instruments and devices do not provide any haptic feedback or additional imaging to visualize hidden structures. In rectal surgery, correct identification of anatomical structures and their boundaries and preparation in thin dissection planes is crucial given the strong association with both local recurrence risk and the risk of incontinence and impaired sexual function [1–4]. In this work, we explored two major computer vision tasks in the context of robot-assisted rectal surgery that could serve as a basis for future context-aware guidance: recognition of surgical phases and exact delineation of target structures including organs and dissection areas.

In summary, differentiation of five surgical phases was feasible at an average F1 score of 0.82 of the best-performing model, MSTCN [20]. Segmentation of anatomical structures and dissection areas demonstrated promising results for selected target structures using models based on DeepLabv3 [28]. However, several challenges related to training data, model development and validation, and clinical translation limit the potential to leverage the potential of machine learning-based segmentation in laparoscopy (Figure 4).

In general, the presented observations are in line with previous studies on AI-based temporal and spatial analysis of laparoscopic images [31]. With regard to surgical phase recognition, similar accuracies around 0.8 have been reported for other laparoscopic procedures including cholecystectomy [20], colorectal surgery [32,33], and laparoscopic sleeve gastrectomy [34]. Previous studies investigating segmentation of anatomical structures have mostly focused on classification of the presence of certain organs [15] or rough localization of the detected organs using bounding boxes [35]. A recent preprint [12] describes semantic segmentation of organs based on a large-scale dataset of organ segmentations [13]. This preprint covers six organs that were also analyzed in the present work. Comparing the results of both works, similarly accurate segmentation performances were observed for the abdominal wall, while considerably inferior segmentation performances were observed for the colon, inferior mesenteric vessels, the small intestine, and seminal vesicles. In part, this inferior performance is likely related to a smaller amount of training data. For instance, algorithms for identification of seminal vesicles and the small intestine were trained on comparably small datasets of 336 and 104 annotated images, respectively (Appendix E), while over 1000 annotated images were used to train models in the aforementioned work. Integration of more and diverse – ideally multicentric – training data would likely improve segmentation performance and increase clinical relevance of respective models [36]. One of the main reasons for scarcity of annotated laparoscopic datasets is the high annotation effort required for an integration of clinically meaningful knowledge into a dataset [37]. In this context, standardization of (video) data recording and adaptability of the annotation process to differences in surgical techniques and approaches pose important challenges to consider for future studies using annotated laparoscopic data. Of note, the Society of American Gastrointestinal and Endoscopic Surgeons recently published recommendations for surgical video annotation to increase the amount of publicly available datasets and enable the creation of benchmark AI algorithms with a clinical impact [38]. With respect to image annotation, a recent analysis has revealed that the inclusion of example images substantially improves annotation quality compared with text-only descriptions, and that annotators with a professional background outperform paid crowdworkers [39]. In the present study, both of these quality criteria are fulfilled (Appendix D), yet only one annotation by advanced medical students was available for creation of the ground truth, which is a major limitation of this work.

Technical approaches to reduce the burden of manual annotation have recently gathered attention in the computer vision community. For example, Sestini et al. have established a fully-unsupervised per-frame segmentation model for surgical instruments based on implicit motion information from surgical videos and information on surgical instrument shape [40]. These so-called instrument shape-priors were derived from existing annotated datasets. Given that the structures and planes segmented for the present study are mostly not covered by existing datasets, such approaches are currently difficult to realize for such complex use cases. In addition, organs and dissection planes do not typically convey motion information to the extent that surgical instruments do. Self-supervised learning methods [41] and active learning [42] are potential approaches that could help overcome the burden of manual segmentation even for complex and novel use cases and rather stationary anatomical regions.

Besides organs, this work also explores feasibility of segmentation of dissection areas. Here, segmentation of the avascular dissection plane during mesorectal excision was promising at an average F1 score of 0.54, which is comparable to previous findings on segmentation of loose connective tissue fibers in gastric surgery [43]. In contrast, exact dissection lines could not be successfully segmented. Especially during mesorectal excision, the appearance of embryonal tissue planes and the closely related dissection line at the mesorectal fascia can vary considerably based on neoadjuvant (radio-)therapy and individual factors such as body composition, making identification particularly challenging [44]. Performance variations of segmentation algorithms were moreover related to common limitations of classical computer vision metrics measuring overlap between ground truth and model prediction [45,46]. For example, such metrics are skewed by segment size and shape, which could further contribute to poor segmentation performance in small and tubular structures such as clips and dissection lines.

The limitations of this work are mostly related to the outlined challenges of AI methods in laparoscopy (Figure 4). First, the monocentric study design and single-rater annotation of training data by non-experts considerably restrict generalizability [47]. Second, segmentation of dissection lines and small structures such as metal clips was largely unsuccessful, likely related to diverse appearance and challenges in existing metrics that are considerably influenced by segment shape and size. To pertain real-world image challenges, images with suboptimal image quality because of camera shake and presence of significant amounts of blood or smoke were not excluded; these factors negatively impacted segmentation performance. Last, the exploratory study design does not allow for estimation of the clinical value of the described phase or target structure recognition models. To enhance applicability and clinical value of the system [48], future works will focus on an integration of recognition uncertainty into a graphical user interface displaying target structure segmentations, for instance through Bayesian computation methods and heatmap-like visualizations [49]. For clinical implementation, such a system will need to meet real-time requirements for clinical usability, necessitating exact determination and subsequent minimization of the computation-associated delay to the range of tenths of a second. The existing limitations notwithstanding, this work represents an important addition to the growing body of research on image-based guidance in abdominal surgery, as it significantly expands the horizon of data science-assisted surgery to procedures of higher complexity and technical difficulty.

## 5 Conclusions

In conclusion, the results suggest that surgical phase recognition and segmentation of selected target structures can satisfactorily be implemented in complex minimally-invasive surgical procedures. These capabilities represent essential components of potential future context-aware guidance systems in complex minimally-invasive oncological surgery. On a general level, this work highlights challenges in the implementation of clinically relevant surgical guidance functions, in particular related to training data, model training and evaluation, and clinical translation. This encourages inter-institutional collaboration and improved data accessibility to facilitate data science-based approaches in a diverse range of surgical procedures with varying complexity.

## Data Availability

The datasets generated and analysed during the current study are available from the corresponding author (Prof. Stefanie Speidel) on reasonable request. To gain access, data requestors will need to sign a data access agreement.

## Abbreviations

AI: Artificial Intelligence
CNN: Convolutional neural network
F1: F1 score
IoU: Intersection-over-Union
LM: Lateral mobilization (phase)
ME: Mesorectal excision (phase)
MM: Medial mobilization (phase)
PME: Partial mesorectal excision
SD: Standard deviation
TME: Total mesorectal excision
VD: Vascular dissection (phase)

## Acknowledgments and Funding

FRK, SL, JW, and SS were supported through project funding within the Else Kröner Fresenius Center for Digital Health (EKFZ), Dresden, Germany (project “CoBot”). FRK received funding from the Medical Faculty of the Technical University Dresden within the MedDrive Start program (grant number 60487) and from the Joachim Herz Foundation (Add-On Fellowship for Interdisciplinary Life Science). FMR received a doctoral student scholarship from the *Carus Promotionskolleg* Dresden. Figures were created with BioRender.com. The authors acknowledge excellent project coordination by Dr. Elisabeth Fischermeier and Dr. Grit Krause-Jüttler.

## Author contributions

FRK, SL, JW, MD, and SS conceptualized the study. FRK, MC, FMR, and TN collected and annotated clinical and video data and contributed to data analysis. SL, SK, and SB implemented and trained the neural networks and contributed to data analysis. JK gave substantial clinical input. JW, MD, JF, and SS supervised the project, provided infrastructure and gave important scientific input. FRK drafted the initial manuscript text. All authors reviewed, edited, and approved the final manuscript.

## Code Availability

The developed framework in this current study based on the DeepLabv3 [28] using the PyTorch library [50]. The source code for surgical phase recognition and the source code for image segmentation are publicly available: https://gitlab.com/nct_tso_public/cobot_phase_recognition (phase recognition) and https://gitlab.com/nct_tso_public/cobot-multi-instance-segmentation (structure segmentation).

