## Supplementary Material for "Artificial Intelligence for context-aware surgical guidance in complex robot-assisted oncological procedures: An exploratory feasibility study"

### Appendix

### Appendix A: Patient characteristics

| | Total (n = 57) | Temporal annotation (n = 33) | Vascular Dissection (n = 10) | Medial Mobilization (n = 11) | Lateral Mobilization (n = 10) | Mesorectal Excision (n = 10) |
|---|---|---|---|---|---|---|
| **Age** [years] | 62.4 ± 11.2 | 62.2 ± 10.2 | 62.1 ± 5.2 | 65.6 ± 7.7 | 63.9 ± 5.0 | 62.8 ± 8.1 |
| **BMI** [kg/m²] | 26.5 ± 3.3 | 26.7 ± 2.9 | 26.1 ± 2.1 | 26.7 ± 2.9 | 26.6 ± 2.8 | 26.9 ± 2.0 |
| **Surgery duration** [min] | 414 ± 112 | 415 ± 117 | 444 ± 148 | 410 ± 98 | 466 ± 141 | 377 ± 130 |
| **Sex** | | | | | | |
| Female | 14 (24.6) | 5 (15.2) | 1 (10.0) | 2 (18.2) | 1 (10.0) | 1 (10.0) |
| Male | 43 (75.4) | 28 (84.8) | 9 (90.0) | 9 (81.8) | 9 (90.0) | 9 (90.0) |
| **Indication** | | | | | | |
| CRC | 54 (94.7) | 31 (93.9) | 10 (100.0) | 11 (100.0) | 10 (100.0) | 10 (100.0) |
| Other | 3 (5.3) | 2 (6.1) | 0 | 0 | 0 | 0 |
| **Distance from anocutaneous line** | | | | | | |
| <6 cm | 19 (33.3) | 14 (42.4) | 3 (30.0) | 3 (27.3) | 3 (30.0) | 5 (50.0) |
| 6 - <12 cm | 26 (45.6) | 14 (42.4) | 4 (40.0) | 5 (45.5) | 5 (50.0) | 3 (30.0) |
| ≥12 cm | 12 (21.1) | 5 (15.2) | 3 (30.0) | 3 (27.3) | 2 (20.0) | 2 (20.0) |
| **Tumor stenosis** | | | | | | |
| Yes | 23 (40.4) | 11 (33.3) | 4 (40.0) | 5 (45.5) | 3 (30.0) | 4 (40.0) |
| No | 34 (59.6) | 22 (66.7) | 6 (60.0) | 6 (54.5) | 7 (70.0) | 6 (60.0) |
| **Neoadjuvant irradiation** | | | | | | |
| Yes | 25 (43.9) | 16 (48.5) | 4 (40.0) | 4 (36.4) | 4 (40.0) | 4 (40.0) |
| No | 32 (56.1) | 17 (51.5) | 6 (60.0) | 7 (63.6) | 6 (60.0) | 6 (60.0) |
| **Previous intraabdominal surgery** | | | | | | |
| Yes | 20 (35.1) | 10 (30.3) | 1 (10.0) | 0 | 1 (10.0) | 3 (30.0) |
| No | 37 (64.9) | 23 (69.7) | 9 (90.0) | 11 (100.0) | 9 (90.0) | 7 (70.0) |
| **Surgical resection technique** | | | | | | |
| LAR, TME | 33 (57.9) | 18 (54.5) | 4 (40.0) | 7 (63.6) | 6 (60.0) | 4 (40.0) |
| ISR, TME | 9 (15.8) | 5 (15.2) | 1 (10.0) | 0 | 1 (10.0) | 2 (20.0) |
| APR, TME | 6 (10.5) | 4 (12.1) | 1 (10.0) | 0 | 0 | 2 (20.0) |
| AR, PME | 9 (15.8) | 6 (18.2) | 4 (40.0) | 4 (36.4) | 3 (30.0) | 2 (20.0) |
| **T status** (for rectal cancers, n = 54) | | | | | | |
| pT0 | 4 (7.4) | 2 (6.5) | 0 | 1 (9.1) | 0 | 2 (20.0) |
| pTis | 1 (1.9) | 0 | 0 | 0 | 0 | 0 |
| pT1 | 8 (14.8) | 4 (12.9) | 3 (30.0) | 2 (18.2) | 3 (30.0) | 3 (30.0) |
| pT2 | 14 (25.9) | 9 (29.0) | 2 (20.0) | 1 (9.1) | 2 (20.0) | 3 (30.0) |
| pT3a | 15 (27.8) | 10 (32.3) | 3 (30.0) | 6 (54.5) | 3 (30.0) | 1 (10.0) |
| pT3b | 12 (22.2) | 6 (19.4) | 2 (20.0) | 1 (9.1) | 2 (20.0) | 1 (10.0) |
| **N status** (for rectal cancers, n = 54) | | | | | | |
| pN0 | 32 (59.3) | 14 (45.2) | 4 (40.0) | 4 (36.4) | 4 (40.0) | 5 (50.0) |
| pN1 | 14 (25.9) | 10 (32.3) | 5 (50.0) | 3 (27.3) | 5 (50.0) | 3 (30.0) |
| pN2a | 5 (9.3) | 4 (12.9) | 1 (10.0) | 2 (18.2) | 0 | 1 (10.0) |
| pN2b | 3 (5.6) | 3 (9.7) | 0 | 2 (18.2) | 1 (10.0) | 1 (10.0) |

**Appendix A**: **Patient characteristics.** For age, BMI and surgery duration, mean ± SD are displayed. All other data are presented as total numbers and percentages of the (sub-)cohort. Abbreviations: Abdominoperineal resection (APR), Anterior Resection (AR), Body Mass Index (BMI), Colorectal cancer (CRC), Low anterior resection (LAR), Intersphincteric resection (ISR), Partial mesorectal excision (PME), Total mesorectal excision (TME).

### Appendix B: Annotation protocol summary for temporal annotation of RARR videos

**P1: Preparation and Intraabdominal Orientation**

- Trocar Placement
- Intraabdominal orientation
- Detachment of superficial adhesions on the abdominal wall

**P2: Medial Mobilization of Descending Colon***

- Incision of the mesocolic peritoneum
- Preparation, clipping, and dissection of the inferior mesenteric artery
- Preparation, clipping, and dissection of the inferior mesenteric vein
- Separation of the mesocolon and Gerota's fascia from medial to lateral

**P3: Lateral Mobilization of Descending Colon***

- Incision along the confluence of the parietal peritoneum of the left-lateral abdominal wall with the colonic visceral peritoneum (*white line of Toldt*)
- Separation of the mesocolon and Gerota's fascia from lateral to medial

**P4: Mesorectal Excision**

- Peritoneal incision superior to peritoneal fold
- Circular mesorectal dissection
- Linear stapling of the rectum

**P5: Extraabdominal Preparation**

- Pfannenstiel incision
- Open preparation and dissection of the descending colon
- Placement of circular stapler head
- Transverse coloplasty
- Temporary closure of the Pfannenstiel incision

**P6: Intraabdominal Preparation of Anastomosis****

- Circular Stapling of the (neo-)rectum
- Water Probe

**P7: Ileostomy****

**P8: Closure****

* Medial and lateral mobilization of the descending colon can be repeated several times until full mobility of the descending colon was achieved.

** Due to inconsistent recording of P6-P8, these phases were not considered for phase identification in this study.

### Appendix C: Selection criteria of videos for semantic segmentation of target structures

| Phase | Target structure | Specifications |
|---|---|---|
| VD | Inferior Mesenteric Artery | Visually delimitable artery wall, minimal residual (fatty) tissue covering was included in the segment |
| | Inferior Mesenteric Vein | Visually delimitable vein wall; minimal residual (fatty) tissue covering was included in the segment |
| | Plastic Clip | *Weck Hem-o-lok® Polymer Locking Ligation System* |
| | Metal Clip | *AESCULAP Challenger® Ti Ligation-Clips* |
| MM | Gerota's Fascia | Visually delimitable fascia, minimal residual (fatty) tissue or small amounts of blood were included in the segment |
| | Mesocolon | Mesocolon, which was separated from Gerota's fascia, colon wall was not included in the segment |
| | Dissection Line (MM) | Visually delimitable line between *Gerota's Fascia* and *Mesocolon* |
| LM | Abdominal Wall | Non-altered abdominal wall; detached adhesions or incisions were excluded from the segment |
| | Adhesion | Adhesion of epiploic appendices or colon to the abdominal wall |
| | Colon | Colon wall; minimal residual (fatty) tissue or small amounts of blood were included in the segment |
| | Fat | All surrounding fatty tissue (e.g. epiploic appendices and greater omentum) |
| | Small Intestine | Outer wall of the small intestine; minimal residual (fatty) tissue or small amounts of blood were included in the segment |
| ME | Mesorectum | Mesorectal envelope, encased by the mesorectal fascia |
| | Dissection Plane (ME) | Avascular plane between the parietal pelvic fascia and the mesorectal fascia ("holy plane") |
| | Dissection Line (ME) | Dissection line at the mesorectal margin of the *Dissection Plane (ME)* |
| | Seminal Vesicles | Visually delimitable white gland structure ventral to the mesorectum |

**Appendix D: Annotation protocol for semantic segmentation of anatomical structures, tissue structures, and dissection planes**

### General annotation guidelines

- Mark only intact and clearly visible wall or parenchyma of respective structures
- Avoid marking
  - Areas substantially covered with smoke/blood/fat
  - Areas not properly visible due to soiling of the laparoscope
  - Large alimentary vessels (include small vessels in the segmentation)
  - Devices and materials such as surgical instruments, compresses, needles, threads
  - Any areas outside the image margins
- Mark structures whenever it is possible to recognize them in an area that is
  - dark
  - slightly covered with smoke, blood, tissue etc.
  - small: sometimes, parts of the structure are visible in tiny areas, e.g. in instrument slots
- Note that a structure may be visible in several small areas in a single image

### Phase-specific annotation guidelines

### (1) Vascular Dissection (VD)

Typically, the Inferior Mesenteric Artery (IMA) is exposed, clipped, and dissected following incision of the visceral peritoneum. Subsequently, the Inferior Mesenteric Vein (IMV) is exposed at the inferior pancreatic margin, clipped, and dissected. Rarely, the IMV is dissected prior to the IMA.

- **Inferior Mesenteric Artery (IMA)**: Mark in images where the vessel wall of the IMA is visible
  - Typical color: white to light pink, often small alimentary vessels are visible
  - Risk of confusion with veins: veins are usually more bluish in color and appear softer
  - When in doubt, it can be helpful to identify the artery at the moment of clipping in order to recognize it in images from earlier phases of the surgery
- **Inferior Mesenteric Vein (IMV)**: Mark in images where the vessel wall of the IMV is visible
  - Bluish color
  - Risk of confusion with larger retroperitoneal veins (i.e. the testicular vein)
  - Risk of confusion with arteries: arteries usually have a whiter color and appear stiffer
  - When in doubt, it can be helpful to identify the vein at the moment of clipping in order to recognize it in images from earlier phases of the surgery
- **Plastic Clip**: *Weck Hem-o-lok® Polymer Locking Ligation System*
  - White plastic clip
  - Mark whenever the clip is visible in the frame, even when it is still anchored in the instrument or attached to the vessel
- **Metal Clip**: *AESULAP Challenger® Ti Ligation-Clips*
  - Narrow metallic clip
  - Mark whenever the clip is visible in the frame, even when it is still anchored in the instrument or attached to the vessel

**Example annotations: Vascular Dissection (VD)**

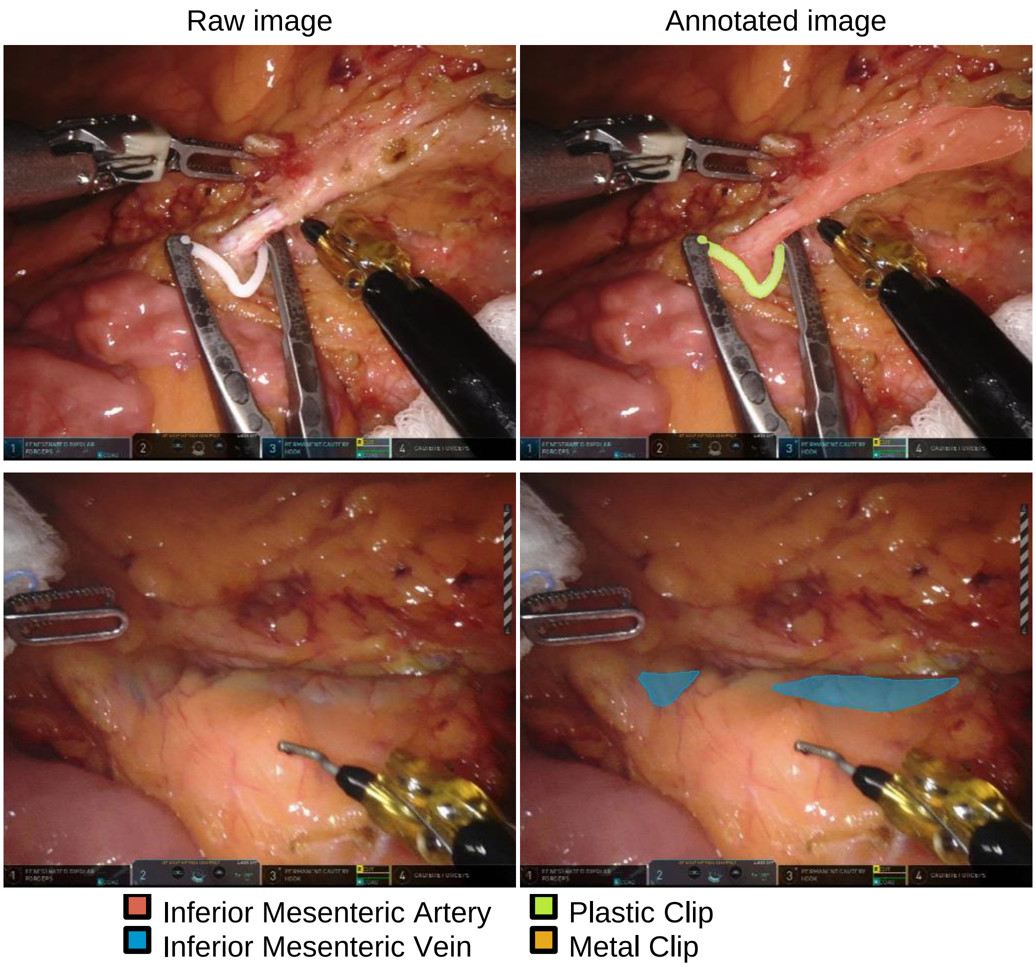

### (2) Medial Mobilization of Descending Colon (MM)

In this phase, the mesocolon is separated from Gerota's fascia to mobilize the descending colon.

- **Gerota's Fascia**: Mark Gerota's fascia (even if there are small blood or fat deposits)
  - Typical color: pink-yellowish, with surface reflections
  - Dorsal of the mesocolon
  - During mobilization, connective tissue between mesocolon and Gerota's fascia is separated
- **Mesocolon**: Mesocolon, when detached from Gerota's fascia
  - Typical color: dark yellow (mesocolic fat)
  - Ventral of Gerota's fascia
  - During mobilization, connective tissue between mesocolon and Gerota's fascia is separated
- **Dissection Line (MM)**: Dissection line between mesocolon and Gerota's fascia
  - Fine line of connective tissue between mesocolon and Gerota's fascia, which is visible when light tension is applied to both structures

**Example annotations: Medial Mobilization of Descending Colon (MM)**

Raw image Annotated image

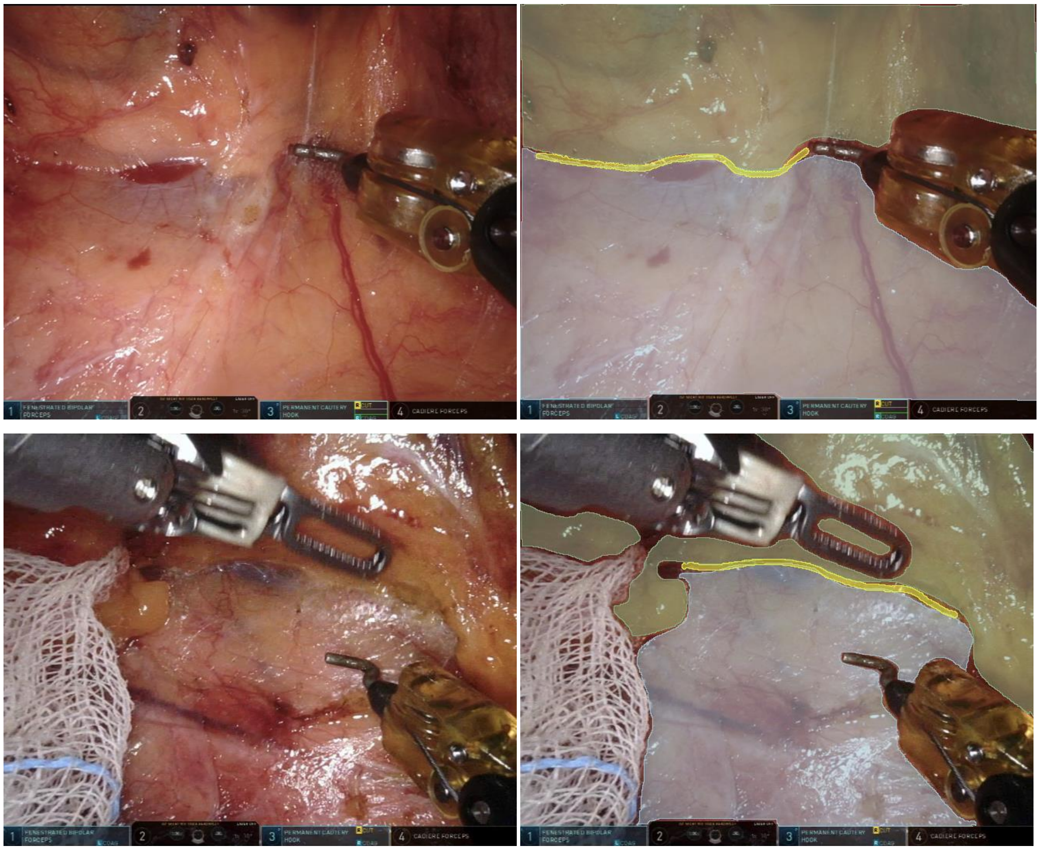

Gerota's Fascia
Mesocolon
Dissection Line (MM)

### (3) Lateral Mobilization of Descending Colon (LM)

In this phase, the descending colon is detached from the lateral abdominal wall. This phase may occur several times during the operation.

- **Abdominal Wall**: Intact lateral abdominal wall
  - Only mark fatty tissue that is located within the abdominal wall layers (i.e. areas covered with parietal peritoneum); omit superficial fat appendages on the abdominal wall
  - Pay close attention to the dorsal borders of the abdominal wall segment when the colon is still attached to the abdominal wall
  - Folds of the abdominal wall (e.g. plica umbilicalis) should be marked
- **Adhesion**: Areas where the colon or the epiploic appendices adhere to the abdominal wall
  - Whitish and fascia-like connective tissue
  - Often seen in several places in one image
- **Colon**: Intact colon wall
  - Risk of confusion with small intestine: colon has epiploic appendices, haustra, taeniae
  - Colon is often attached to the abdominal wall in the images

  - Avoid marking areas with fat appendages, for example appendices epiploicae or remnants after the separation of the colon and the greater omentum
- **Fat**: Fatty tissue (i.e. epiploic appendices and greater omentum)
  - Typical color: yellowish, with surface reflections
- **Small Intestine**: Intact small intestine wall
  - Risk of confusion with colon: small bowel usually is narrower, lighter in color and smoother than colon; small bowel does not have appendices epiploicae, haustra, taeniae; peristalsis can resemble haustra

**Example annotations: Lateral Mobilization of Descending Colon (LM)**

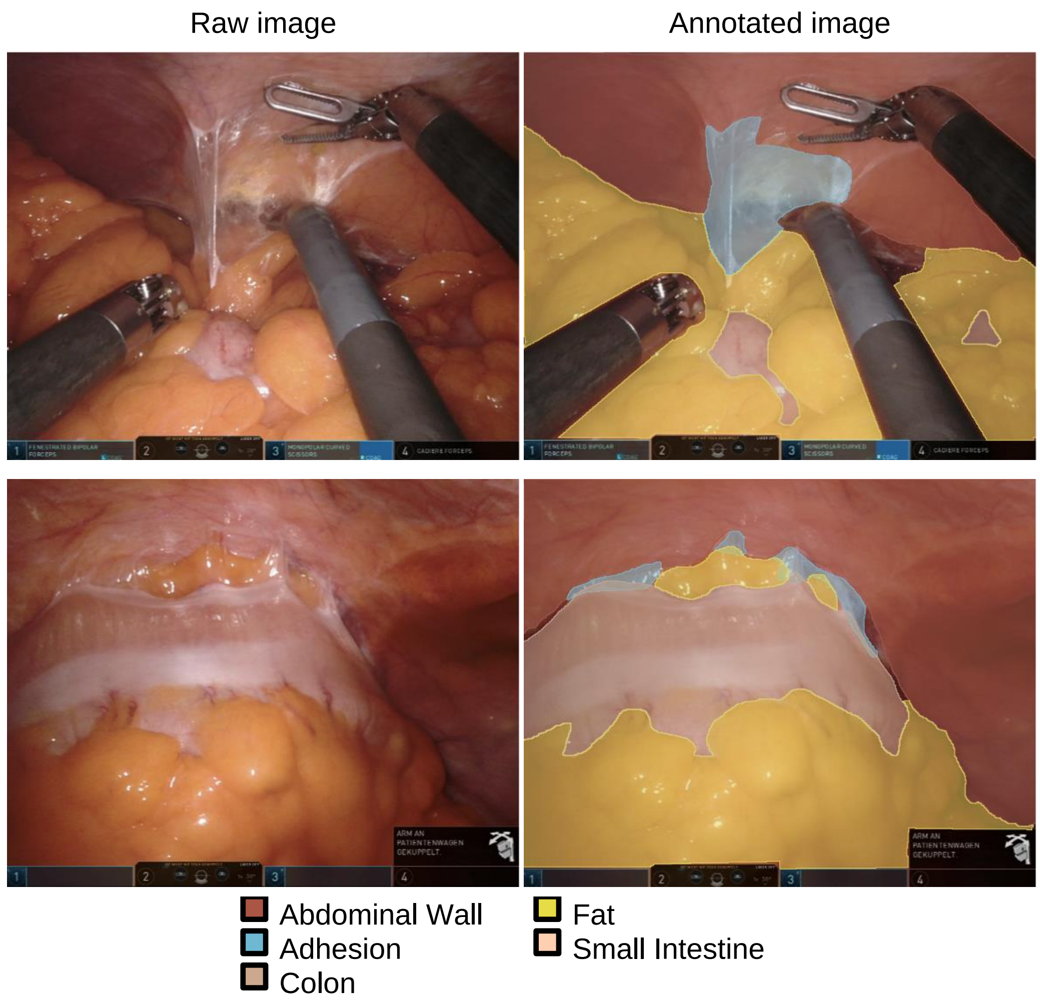

### (4) Mesorectal Excision (ME)

Dorsal, lateral and ventral dissection of the rectum within the mesorectal fascia, usually until the pelvic floor is reached.

- **(Meso-)Rectum**: Mesorectum and/or fatty layers of the rectum that have been detached from the parietal pelvic fascia
  - Typical color: yellow-pink

  - Given that tension into all directions is applied to the mesorectum, the mesorectum is usually located at the outside of the image
- **Dissection Plane (ME)**: Fascia structure surrounding the mesorectum, consisting of Denonvillier's fascia, the parietal pelvic fascia and Waldeyer's fascia
  - Whitish and fascia-like tissue structure
  - Annotate the entire area where the whitish fascia is visible
  - May be visible in different areas in the image based on the direction into which tension is applied to the (meso-)rectum
- **Dissection Line (ME)** between the Dissection Plane (ME) and the mesorectal fascia
  - Variable visibility depending on individual factors (i.e. radiotherapy, constitution)
  - May be visible in different areas in the image based on the direction into which tension is applied to the (meso-)rectum
- **Seminal Vesicles**
  - Color: white to pink
  - Initially, sometimes only very small areas are visible. Skipping a few images ahead can help detect the correct area

**Example annotations: Mesorectal Excision (ME)**

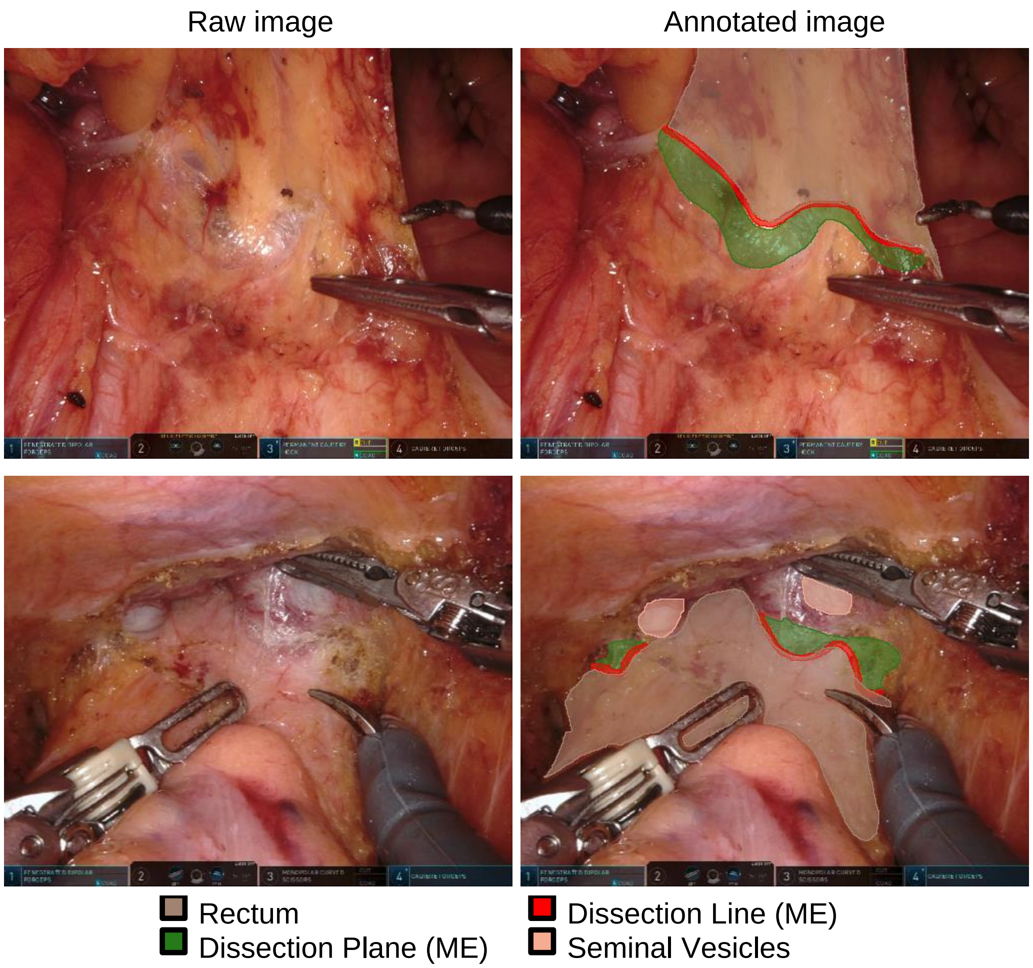

### Appendix E: Distribution of annotated video frames with respect to surgery phases and individual target structures

| Phase | Target structure | Annotated images per target structure | Total annotated images |
|---|---|---|---|
| **VD** | Inferior Mesenteric Artery<br>Inferior Mesenteric Vein<br>Plastic Clip<br>Metal Clip | 1719<br>1510<br>683<br>760 | 3350 |
| **MM** | Gerota's Fascia<br>Mesocolon<br>Dissection Line (MM) | 1847<br>1852<br>1289 | 1880 |
| **LM** | Abdominal Wall<br>Adhesion<br>Colon<br>Fat<br>Small Intestine | 1084<br>782<br>906<br>1038<br>104 | 1086 |
| **ME** | Dissection Plane (ME)<br>Dissection Line (ME)<br>Rectum<br>Seminal Vesicles | 2428<br>2326<br>2472<br>336 | 2721 |
| **Total** | | | 9037 |

**Appendix E: Distribution of annotated video frames with respect to surgery phases and individual target structures.** Table displays the number of annotated video frames for training of phase-specific semantic segmentation algorithms and presence of individual target structures in the annotated image frames. Abbreviations: Lateral mobilization (LM), Medial mobilization (MM), Mesorectal excision (ME), Vascular dissection (VD).

### Appendix F: Summary of performance metrics for recognition of surgical phases using a ResNet50 backbone with different pre-training configurations

| Backbone | Pre-training | F1 score | Accuracy |
|---|---|---|---|
| ResNet50 | ImageNet | 0.683 ± 0.013 | 0.739 ± 0.021 |
| ResNet50 | ImageNet + RARR videos (self-supervised) | 0.680 ± 0.010 | 0.737 ± 0.016 |

**Appendix F: Summary of performance metrics for recognition of surgical phases using a ResNet50 backbone with vs. without self-supervised pre-training.** For each metric, mean and standard deviation are displayed.

### Appendix G: Distribution of phase durations

| Phase | Phase duration (including zero-length phases) [seconds] | Phase duration (excluding zero-length phases) [seconds] |
|---|---|---|
| P1: Preparation and intraabdominal orientation | 1394 ±1468 | 1769 ± 1439 |
| P2: Medial Mobilization of Descending Colon | 5337 ± 2121 | 5504 ± 1929 |
| P3: Lateral Mobilization of Descending Colon | 1589 ± 1196 | 1691 ± 1161 |
| P4: Mesorectal Excision | 7851 ± 5281 | 8357 ± 5045 |
| P5: Extraabdominal Preparation | 2020 ± 4302 | 4445 ± 5471 |

Average total video duration (P1 – P5): 18191 seconds

**Appendix G: Distribution of phase durations in 33 temporally annotated full-length surgery videos.** For each metric, mean and standard deviation are displayed.

### Appendix H: Distribution of segment sizes

| Phase | Target structure | Number of pixels per image | % pixels per image |
|---|---|---|---|
| VD | Inferior Mesenteric Artery | 26816 ± 21038 | 4.09 ± 3.21 |
| | Inferior Mesenteric Vein | 12355 ± 13802 | 1.89 ± 2.11 |
| | Plastic Clip | 3215 ± 4474 | 0.49 ± 0.68 |
| | Metal Clip | 2150 ± 2029 | 0.33 ± 0.31 |
| MM | Gerota’s Fascia | 181876 ± 91135 | 27.75 ± 13.91 |
| | Mesocolon | 179514 ± 91880 | 27.39 ± 14.02 |
| | Dissection Line (MM) | 3617 ± 2420 | 0.55 ± 0.37 |
| LM | Abdominal Wall | 184691 ± 76612 | 28.18 ± 11.69 |
| | Adhesion | 20820 ± 22629 | 3.18 ± 3.45 |
| | Colon | 70684 ± 54896 | 10.79 ± 8.38 |
| | Fat | 157900 ± 91381 | 24.09 ± 13.94 |
| | Small Intestine | 65077 ± 48802 | 9.93 ± 7.45 |
| ME | Dissection Plane (ME) | 22616 ± 29901 | 3.45 ± 4.56 |
| | Dissection Line (ME) | 2419 ± 1646 | 0.37 ± 0.25 |
| | Rectum | 109005 ± 68442 | 16.63 ± 10.44 |
| | Seminal Vesicles | 14751 ± 18769 | 2.25 ± 2.86 |

**Appendix H: Distribution of segment sizes.** For each metric, mean and standard deviation are displayed.

### Appendix I: Summary of performance metrics for phase-specific semantic segmentation of target structures and dissection areas (calculated over metric means per surgery)

| Phase | Target Structure | F1 score | IoU | Precision | Recall | Specificity |
|---|---|---|---|---|---|---|
| VD | Inferior Mesenteric Artery | 0.26 ± 0.13 | 0.19 ± 0.10 | 0.42 ± 0.20 | 0.34 ± 0.14 | 1.00 ± 0.00 |
| | Inferior Mesenteric Vein | 0.31 ± 0.17 | 0.24 ± 0.14 | 0.44 ± 0.12 | 0.44 ± 0.22 | 1.00 ± 0.00 |
| | Plastic Clip | 0.29 ± 0.27 | 0.24 ± 0.23 | 0.55 ± 0.29 | 0.42 ± 0.41 | 1.00 ± 0.00 |
| | Metal Clip | 0.14 ± 0.11 | 0.09 ± 0.08 | 0.15 ± 0.13 | 0.58 ± 0.24 | 1.00 ± 0.00 |
| MM | Gerota’s Fascia | 0.84 ± 0.03 | 0.75 ± 0.03 | 0.82 ± 0.04 | 0.90 ± 0.03 | 0.94 ± 0.02 |
| | Mesocolon | 0.79 ± 0.04 | 0.68 ± 0.04 | 0.79 ± 0.05 | 0.84 ± 0.04 | 0.94 ± 0.01 |
| | Dissection Line (MM) | 0.21 ± 0.05 | 0.13 ± 0.03 | 0.21 ± 0.07 | 0.39 ± 0.07 | 1.00 ± 0.00 |
| LM | Abdominal Wall | 0.88 ± 0.03 | 0.80 ± 0.04 | 0.89 ± 0.03 | 0.89 ± 0.04 | 0.96 ± 0.01 |
| | Adhesion | 0.32 ± 0.12 | 0.22 ± 0.09 | 0.36 ± 0.12 | 0.45 ± 0.11 | 0.99 ± 0.01 |
| | Colon | 0.61 ± 0.08 | 0.52 ± 0.07 | 0.66 ± 0.11 | 0.75 ± 0.09 | 0.98 ± 0.01 |
| | Fat | 0.79 ± 0.05 | 0.70 ± 0.05 | 0.78 ± 0.05 | 0.87 ± 0.05 | 0.95 ± 0.01 |
| | Small Intestine | 0.20 ± 0.18 | 0.17 ± 0.16 | 0.29 ± 0.25 | 0.43 ± 0.31 | 1.00 ± 0.00 |
| ME | Rectum | 0.54 ± 0.08 | 0.44 ± 0.07 | 0.59 ± 0.09 | 0.63 ± 0.08 | 0.94 ± 0.02 |
| | Dissection Plane (ME) | 0.50 ± 0.07 | 0.37 ± 0.06 | 0.52 ± 0.07 | 0.63 ± 0.06 | 0.99 ± 0.01 |
| | Dissection Line (ME) | 0.17 ± 0.03 | 0.10 ± 0.02 | 0.17 ± 0.02 | 0.26 ± 0.07 | 1.00 ± 0.00 |
| | Seminal Vesicles | 0.16 ± 0.11 | 0.12 ± 0.09 | 0.29 ± 0.17 | 0.36 ± 0.21 | 1.00 ± 0.00 |

**Appendix I: Summary of performance metrics for phase-specific semantic segmentation of target structures and dissection areas.** For each metric, mean and standard deviation are displayed. Means and standard deviations were calculated over metric means per surgery. Abbreviations: Intersection over union (IoU), Lateral mobilization (LM), Medial mobilization (MM), Mesorectal excision (ME), Vascular dissection (VD).
